# Evaluating the effectiveness of mindfulness alone compared to exercise and mindfulness on fatigue in women with gynaecology cancer (GEMS): Protocol for a randomised feasibility trial

**DOI:** 10.1101/2022.11.14.22282320

**Authors:** Kairen McCloy, Ciara Hughes, Lynn Dunwoody, Joanne Marley, Ian Cleland, Federico Cruciani, Catherine Saunders, Jackie Gracey

## Abstract

**Background:** In 2020 Globocan reported nearly 1.4 million new cases of gynaecology cancer worldwide. Cancer related fatigue has been identified as a symptom that can be present for gynaecology cancer patients many years after treatment. The current evidence around the management of this symptom suggests that exercise has the most positive outcome. However, some ambiguity remains around the evidence and whether it can address all areas of fatigue effectively. More recently, other interventions such as mindfulness have begun to show a favourable response to the management of symptoms for cancer patients. To date there has been little research that explores the feasibility of using both these interventions together in a gynaecology cancer population. This study aims to explore the feasibility of delivering an intervention that involves mindfulness and mindfulness and exercise and will explore the effect of this on fatigue, sleep, mood and quality of life.

**Methods/Design:** This randomised control trial will assess the interventions outcomes using a pre and post design and will also include a qualitative process evaluation. Participants will be randomised into one of 2 groups. One group will undertake mindfulness only and the other group will complete exercise and mindfulness. Both groups will use a mobile application to complete these interventions over 8 weeks. The mobile app will be tailored to reflect the group the participants have drawn during randomisation. Self-reported questionnaire data will be assessed at baseline prior to commencing intervention and at post intervention. Feasibility will be assessed through recruitment, adherence, retention and attrition. Acceptability and participant perspective of participation (process evaluation), will be explored using focus groups.

**Discussion:** This trial will hope to evidence and demonstrate that combination of two interventions such as mindfulness and exercise will further improve outcomes of fatigue and wellbeing in gynaecology cancer. The results of this study will be used to assess (i) the feasibility to deliver this type of intervention to this population of cancer patients using a digital platform; (ii) assist this group of women diagnosed with cancer to manage fatigue and other symptoms of sleep, mood and impact their quality of life.

**Trial registration:** NCT05561413

## Introduction

Gynaecology cancer encompasses 5 main types, ovarian, endometrial, cervical, vaginal, and vulva [1,2]. In the United Kingdom (UK) 21,493 new cases of gynaecological cancer are diagnosed every year [2]. Treatment modalities such as surgery, chemotherapy or radiotherapy can be effective in managing and treating these cancers but may lead to unwanted long term side effects. One side effect is fatigue, the prevalence of which, for all types of cancer, was identified as 52%. For the gynaecology cancer population, this varied between 17-33%, with many women experiencing cancer related fatigue (CRF) years after treatment [3,4]. CRF has been described not only as a physical sensation, but also has emotional and cognitive symptoms suggesting that it is multi-dimensional [5]. To date, few studies have evaluated interventions that target all dimensions of fatigue, suggesting a multi-modal approach that addresses all aspects is required.

### Mechanisms

Risk factors for CRF, include depression and insomnia suggesting that the presence of these are not only correlated but may also have an impact on the levels and severity of CRF [6]. Indeed, the term ‘cluster symptom’ has recently been adopted to describe this and the most common symptoms in this cluster have been identified as depression, insomnia, and fatigue [7]. This suggests that managing fatigue may require the incorporation of interventions that deal with the multiple symptoms of depression, insomnia, and fatigue collectively [8,9].

### Existing Knowledge

Research indicates that exercise can have a positive effects on CRF [10–15]. However, the effect of exercise on CRF and quality of life (QoL) remains unclear [1,16] with some studies having reported positive outcomes and others demonstrating no change [15,17,18]. This ambiguity may suggest that exercise alone may not be enough to ameliorate CRF and improve QoL and a multidimensional approach may be required.

Despite the evidence of the positive effect of exercise on CRF, adherence to exercise remains a problem, with less than 20% of women with ovarian cancer exercising regularly following treatment [19]. Barriers to exercise include time constraints, cost, weather, side effects of treatment or exercise such as fatigue, social aspects, not being aware of physical activity guidelines, fatigue, and psychological barriers, such as motivation [20,21]. In contrast, the facilitators to exercise identified within the literature were improved physical and mental wellbeing, decreased feelings of stress, enjoyment, satisfaction, improved levels of CRF and control over health [22–24].

Research indicates that mindfulness can have a positive effects on both CRF and QoL [25–29]. Mindfulness involves being intentionally aware of the present moment and doing this without judgement. Through practice participants develop an awareness of current emotions and thoughts with compassion and kindness which in turn will lead to better control of cognition, emotion and behaviour [30]. However, some of the effect sizes for mindfulness on psychological and physical outcomes such as depression, anxiety, fatigue were small, and did not reach minimal clinical significance. Despite this, it seems that mindfulness had positive effects on psychological distress, which is defined as an emotion that is not pleasant [31–33]. This is of importance as psychological distress and particularly symptoms of anxiety, depression, sleep, stress and quality of life have been shown to improve following the introduction of mindfulness [34–36]. Some of these symptoms are also present within the ‘cluster symptoms’ previously identified suggesting that there may also be an improvement in CRF alongside these symptoms.

Digital interventions for health have grown in number with over 50,000 medical or healthcare available on Google Play or the Apple Apps Store [37]. For cancer patients apps can have various aims including to provide education, identify and mange symptoms, lifestyle promotion and functional exercise [38]. A recent review identified how digital and interactive health interventions had a positive effect on physical and psychological symptoms on women with breast cancer. This review identified interventions such as music therapy, physical training such as walking, swimming, resistance training, dance, support and education and how they were delivered through a digital platform successfully and showed a favourable response [39]. Mindfulness when delivered digitally has also been shown to be feasible and result in a positive outcome [25,40,41]. Suggesting that the delivery of various interventions for cancer patients can be achieved remotely increasing the scale and inclusivity of a wider population.

### Need for a trial

The majority of studies that involve exercise or mindfulness for the management of CRF and psychological distress are in the breast cancer population making it difficult to generalise findings to other cancer populations [42,43]. Both exercise and mindfulness have individually as interventions demonstrated positive effects on CRF, anxiety, depression, and sleep. Whether mindfulness and particularly adding mindfulness to an exercise regime for managing fatigue in women with gynaecology cancer is of benefit, is yet to be explored. Therefore, the aim of this study is to assess how feasible and acceptable the digital delivery of interventions of, mindfulness, and exercise, will be and if there is any effect on CRF for those with gynaecology cancer.

### Theoretical Model

It has been suggested that for the successful development and implementation of complex interventions a theoretical model should be used [44]. For the interventions within this study the COM-B model [45] was used as to support engagement in exercise and/or mindfulness. This model denotes that for behaviour to occur, three conditions must be met, the individual must have the Capability (both physical and psychological), the Opportunity (social and environmental) and the Motivation (reflective and automatic) to engage in the desired behaviour(s). The COM-B model is a hub that sits within the Behaviour Change Wheel (BCW), which was synthesised from previous frameworks and enhances the development of behavioural interventions [46,47]. The second layer of the BCW includes nine intervention functions (education, persuasion, incentivisation, coercion, training, enablement, modelling, environment restructuring and restrictions), which are linked to the 93 behaviour change techniques (BCTs), as described in the Behaviour Change Techniques version 2 [48]. BCTs are the smallest active ingredients of an intervention, documenting them in a standardised way can aid intervention description and replication [49]. The delivery of intervention functions is supported by the outer layer of the BCW, which includes policy categories of environment/social planning; communication/marketing, fiscal measures, legislation, regulation, service provision and guidelines. The interventions within this study used a range of intervention functions in combination which may then impact the constructs of capability, opportunity and motivation and were linked to appropriate BCTs (Box 1).

Box 1 Description of intervention functions* and behaviour change techniques^†^ used in the interventions

**Exercise Intervention Capability**

Physical Capability

➢*Training*: goal setting, reviewing progress and resetting goals, refining goals and (BCT group 1), guidance with the app and through phone calls on increasing difficulty of task and habit forming (BCT Group 8)

➢*Education:* videos and written instructions through app will guide participation to perform the activities (BCT group 6).

Psychological capability

➢*Training*: identifying problems and problem solving, problem solving strategies (BCT group 1), setting regular time for activities, getting environment ready in advance building habits (BCT group 8).

➢*Education*: within the mobile app written and audio information on the health benefits and consequences (BCT group 5).

**Opportunity**

Physical opportunity

➢*Enablement:* goal setting will be through the mobile app and via weekly phone contact (BCT group 1) and feedback and monitoring will be done via the weekly contact phone call and through the portal where reported data will be viewed by the research team.

Social opportunity

➢*Environmental restructuring:* family support to enable participants to complete activities or buddy up with a family or friend to complete intervention (BCT group3).

**Motivation**

Automatic

➢*Incentivisation*: given through feedback as praise and encouragement of activities and completion of goals (group 2). Encouraging self-belief through giving feedback at how well participants are progressing (BCT group 15)

Reflective

➢*Education:* through app the information on the health consequences of exercise how may improve symptoms of fatigue, sleep, mood. The pros and cons of the activities (BCT group 5 and 9)

**Mindfulness Intervention Capability**

➢*Education:* audio and written instructions through app will guide participation to perform the activities (BCT group 6) and within the mobile app written and audio information on the health benefits and consequences (BCT group 5).

➢*Training:* formal practices within the app along with weekly contact to enable an action plan to perform the practice (BCT group 1), enabling the regular practice which will then lead to habit forming (BCT group 8).

**Opportunity**

Social opportunity

➢*Environmental restructuring:* family support to enable participants to complete activities along with finding an environment that is quite and suitable for practice (BCT group3).

**Motivation**

Automatic

➢*Incentivisation*: given through feedback as praise and encouragement of activities and completion of goals (group 2).

Reflective

*Education:* through app the information on the health consequences of exercise how may improve symptoms of fatigue, sleep, mood. The pros and cons of the activities (BCT group 5 and 9)

*Nine intervention functions identified in the Behaviour Change Wheel framework: education, persuasion, incentivisation, coercion, training, restriction, environmental restructuring, modelling, enablement. [45]

^†^Behaviour change techniques as grouped in BCT Taxonomy (v1): 93 hierarchically-clustered techniques [84]

## Methods and Design

### Primary Aim

To explore the feasibility of conducting an 8-week mindfulness and home-based walking and strength training compared to a mindfulness only programme for the management of CRF in women with gynaecology cancer delivered through an online application. The following objectives will assess feasibility outcomes:

1. To assess participant intervention adherence, study retention and attrition rate.
2. Explore the acceptability of using an mobile application for the delivery and data collection of the exercise and mindfulness programme.
3. Explore the perspectives of participants concerning satisfaction (e.g. intervention and delivery) perceived value, barriers and facilitators of the digital programme.

### Hypotheses of efficacy

Compared to active comparator of mindfulness, the combined intervention of mindfulness and exercise will be superior in reducing fatigue and improving mood, sleep and quality of life.

### Secondary aim

To evaluate the effect of an 8 week exercise and mindfulness programme on cancer related fatigue as a primary clinical outcome and the effect on anxiety, depression, sleep and QoL as a secondary clinical outcome in an digital setting.

### Study design

The Gynaecology Exercise Mindfulness Study (GEMS) will be a randomised controlled open label study. Blinding to this type of study and intervention is not possible, as outcomes are self-reported and in order to complete the programme participants will need to be aware of the arm drawn. Randomisation will be performed as block randomisation using 1:1 ratio.

Participants will be randomised into the intervention group, mindfulness and exercise, or the active comparator group mindfulness only. This will allow assessment of effectiveness of the intervention for both groups on the outcome measures.

Following the intervention participants will be invited to join an online focus groups which will form the qualitative evaluation.

### Study setting

The GEMS trial will be conducted remotely through a mobile application interface allowing recruitment across the UK and asynchronous delivery of the intervention. This app is a protype that has been developed in collaboration with the School of Computing at Ulster University. Pilot testing of this app has taken place with the 5 user testers and adaptations to the app have been made based on this.

### Measurements

Primary measures include the feasibility and acceptability of the study and will be assessed in line with the MRC guidance on developing and evaluating complex interventions [44]. Participant reported outcome measures (PROMs) will be collected for secondary outcomes through a battery of 6 validated questionnaires described below. Estimated completion time of questionnaires is 15 to 20 minutes. See Fig. 1 for an overview of schedule of enrolment, interventions and assessments. At baseline participants will also complete demographic information such as age, cancer type, education, income, and education

**Fig. 1.**
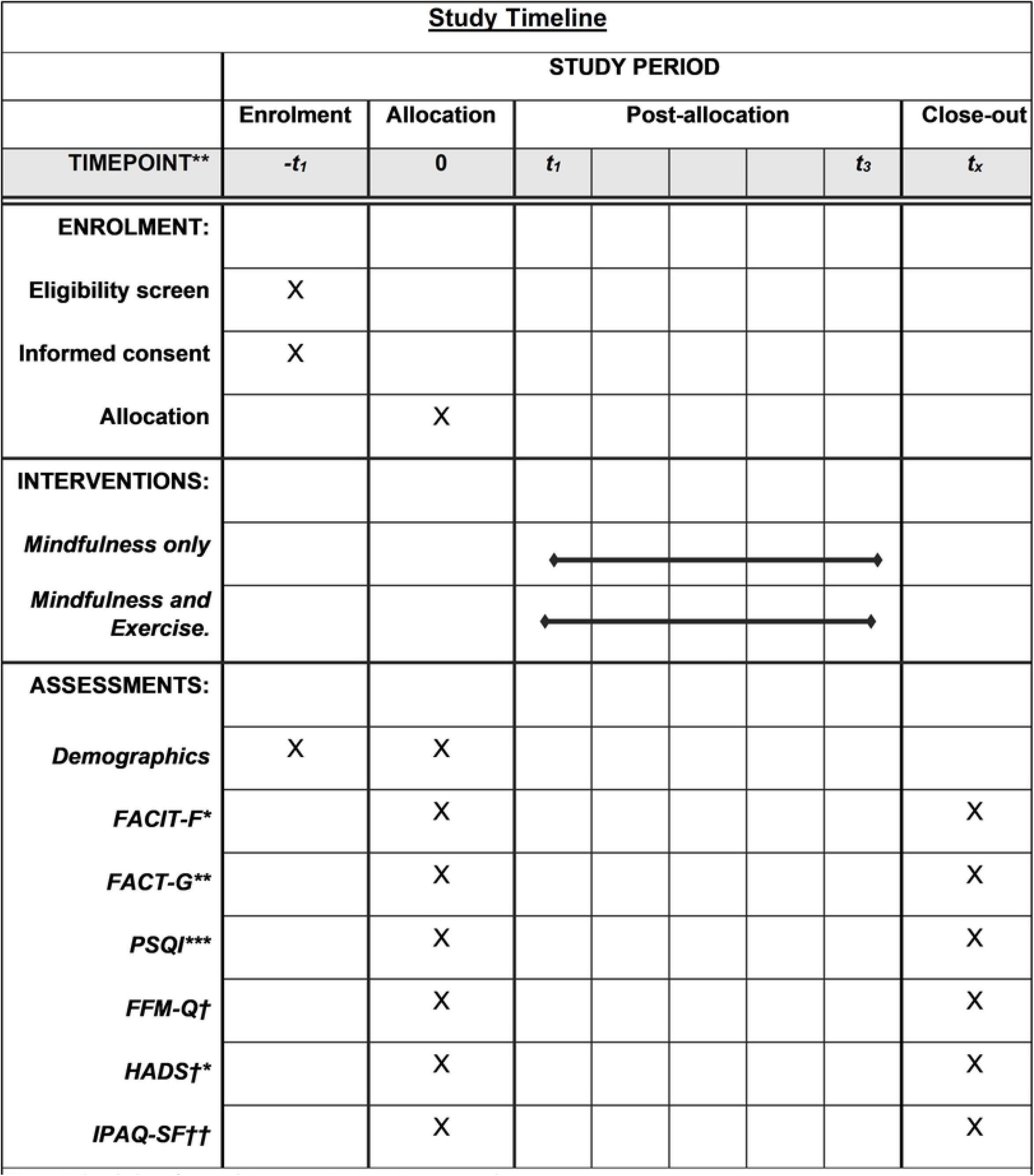
Schedule of enrolment, interventions, and assessment. * The Functional Assessment of Chronic Therapy-Fatigue (FACIT-F) ** The Functional Assessment of Cancer Therapy General (FACT-G) *** Pittsburgh Sleep Quality Index (PSQI) †Five Facet Mindfulness Questionnaire Short Form (FFMQ-SF) †*Hospital Anxiety and Depression Score (HADS) ††lnternational physical activity questionnaire short form (IPAR-Q -S

### Primary outcome measures

Adherence:--interventions will run over 8 weeks. The completion of 4 weeks for mindfulness and 30 minutes of home-based exercise weekly for two thirds (67%) of the duration of the trial will be regarded as adherence to the program. These levels of adherence are based on previous studies that have used the same levels [30,50]. This will be assessed through the electronic logs that participants will be encouraged to complete along with the app data information-on frequency and duration of participant engagement with each session.

Attrition:-Following a review of web-based mindfulness intervention studies, it is estimated that the attrition rate will be approximately 20% with 10% to 15% of missing data [51,52].

Time to recruit, recruitment and retention rates will be gathered through administrative data during (pre)screening, enrolment, allocation and follow-up. Reasons for non-participation, intervention/study dropouts will be collected when possible. Acceptability of data collection will be retrieved through missing data within questionnaires.

Acceptability of the programme will be assessed using focus groups this will include participants experience on the virtual delivery and data collection along with their experience and reflections on the exercise and mindfulness components of the intervention.

### Secondary outcome measures

Fatigue:-The Functional Assessment of Chronic Therapy-Fatigue (FACIT-F) subscale is a 13-item questionnaire that uses a 5 point Likert scale and has been validated for use with various types of cancer and treatments [52,53]. A lower score indicates a greater degree of fatigue, with scores less than 20 representing severe fatigue and scores greater than 45 as normal [14]

Quality of life-The Functional Assessment of Cancer Therapy General (FACT-G) is a 28-item scale. It assesses physical, social, emotional and functional wellbeing. [54]. Higher scores indicate better quality of life [30]

Psychological outcomes will be assessed using the Hospital Anxiety and Depression Scale (HADS) [55]. The HADS consists of 14 questions, 7 each for assessing anxiety and depression [56]. A HADS score of greater than 11 is more defining of anxiety and depression, with a reduction of 3 points indicating a clinical improvement [57].

Sleep-The Pittsburgh Sleep Quality Index (PSQI) is a 19 items questionnaire that measures 7 domains of sleep, scored on a 0-3 scale over 1 month [58]. PSQI scores can range from 0-21, higher scores indicate poorer sleep quality [59].

Mindfulness-The 24 item Five Facet Mindfulness Questionnaire Short Form (FFMQ-SF) is a 5 point Likert Scale which assess mindfulness in daily life. It measures 5 sub-scales: observing, describing, acting with awareness, non-judgement of inner experience and non-reacting to inner experience. Higher scores indicate more mindful in everyday life [60,61]

Participants exercise behaviour will be assessed using the International physical activity questionnaire short form (IPAR-Q -SF). This questionnaire has 7 questions which assess the intensity, frequency and duration of exercise that participants engaged in over the last week.

## Data collection and procedure

Data collection and procedure within the study are presented in the flow chart below (Fig 2).

**Fig 2.**
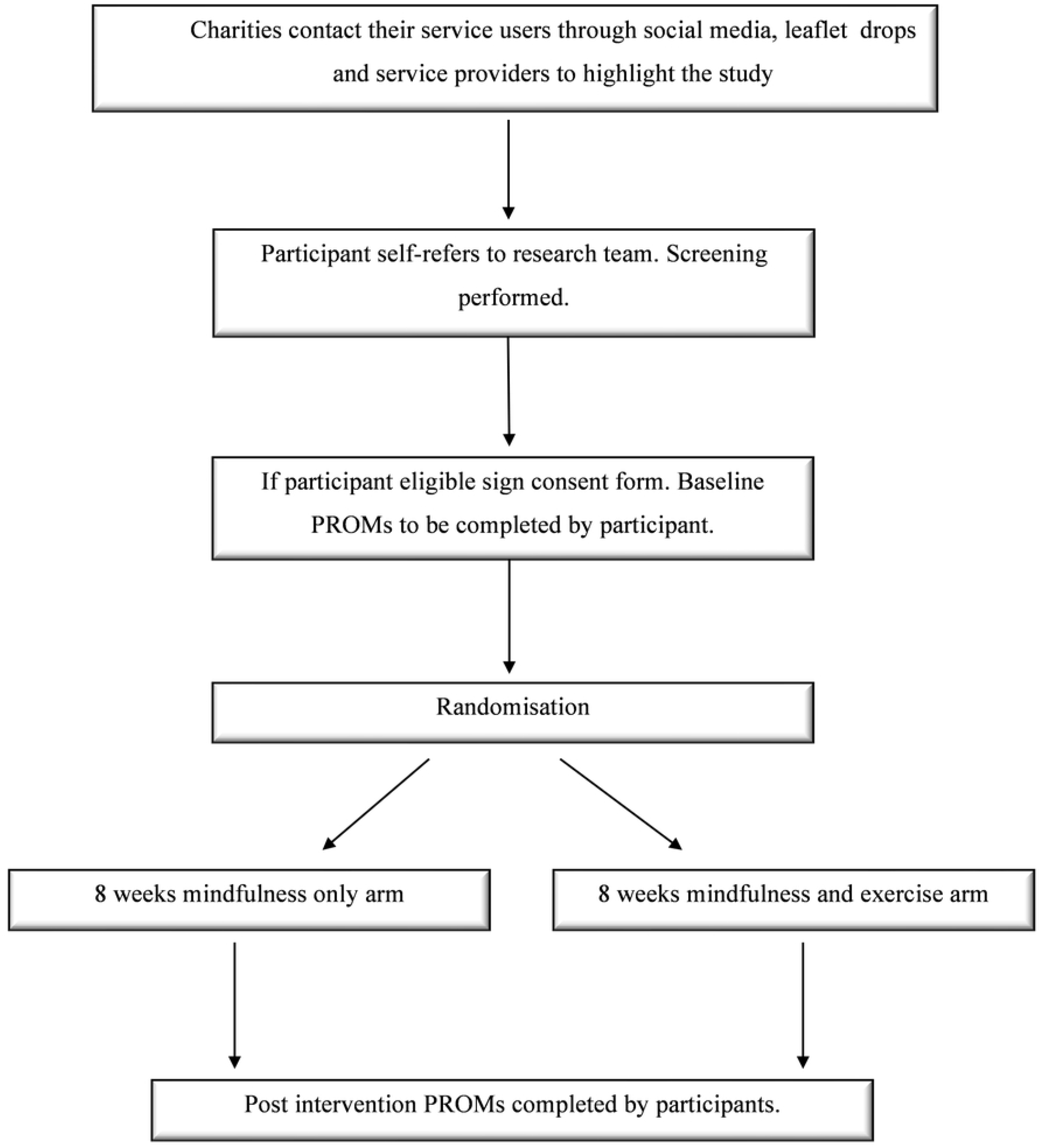
Flow chart of study logistics and data collection.

### Recruitment

Recruitment to the study will be facilitated through cancer charities who will reach out to their service users via social media, forums and groups. The advert will have e-mail contact details for the research team where potential participants can self-refer for further information. When potential participants contact the research team for information the participant information sheet can be e-mailed to them. Those who wish to take part in the study will be screened for eligibility and those who meet the criteria will receive an e-mail link to complete the consent form through Qualtrics© online software. Once consented participants will receive a further Qualtrics© link, containing the online questionnaire (PROMs) that must be filled out (once before randomisation at baseline and then at 8 weeks post intervention).

### Allocation

Participants will be randomly allocated to either group through 1:1 allocation. Block randomisation will be used with fixed block sizes of 10 to ensure an even distribution of groups will be achieved. Study randomisation will be done through Study Randomizer (2017) a web based randomisation service. One member of the research team (KMCC) will carry out all randomisation. Once enrolled all baseline questionnaires (PROMs) have been completed Study Randomizer will be used for allocation to groups [62]. Participants will be notified of group via e-mail.

### Drop out and data retention

If a participant no longer wishes to take part in the study, they can inform the research team of their decision either through e-mail or phone. The study team will contact the participant who has withdrawn either by e-mail or phone to ask if they are willing to provide a reason for withdrawal. In line with institutional requirements, data will be retained for 10 years.

### Inclusion Criteria

Participants must fulfil the following criteria to be eligible for inclusion in the study:

- Women diagnosed with gynaecology cancer
- Over the age of 18
- Still be experiencing fatigue at level 4 or above when assessed on a 10 point single-item scale where 0=‘no fatigue’ and 10=‘greatest possible fatigue’ [63]

### Exclusion Criteria

- Currently actively and regularly practising mindfulness
- Have a confirmed diagnoses of schizophrenia spectrum disorder, bipolar disorder, post-traumatic stress disorder, or risk factors for psychosis (e.g. personality disorder)
- Have an existing medical condition that may inhibit safe participation in the exercise part of the intervention study.

## Interventions

### Exercise Intervention

The exercise component of the study is based around home-based walking and strength and conditioning exercise which have been shown to not only be effective for cancer-related fatigue but also feasible and acceptable [64,65]. Indeed, the research team have demonstrated the positive effects with women who have gynaecology cancer in a previous study and this work will further progress these findings [15,22]. The overall aim of the programme is to achieve a goal of 30 minutes of walking 3 times per week and 2-3 strength sessions, with major muscle groups per week by the end of the 8-week intervention [15,66]. These goals are suggestions, and this programme will use an inductive approach to goal setting which will be decided through collaboration with participants and individual goal setting. A goal setting diary will be provided to participants to aid them to set and monitor goals throughout the study. Alongside this participants can set goals within the mobile app. This approach has been used in previous research conducted by this team in ‘Back on Track’ and ‘EXACT’ trials [67,68].

All materials including audio, video and written information and demonstrations of individual strength exercises, for the study will be delivered through the mobile app which participants can download following randomisation. This programme is delivered asynchronously so participants can commence the programme as soon as they download the app and choose when they wish to complete the interventions. This also allows for rolling recruitment and no requirement for a set number of participants to be recruited and randomised before any participants can commence the study. Participants will be asked to report their activities daily through the app, a daily reminder will be sent to aid reporting. Participants will be contacted by the researcher every week at a mutually agreed time. Contact will be used to assess if there are any issues, set goals for the following week and capture any missing data a standardised questionnaire will be used to assess this. Engagement and usage of the app will be captured, and this data will be analysed feasibility of delivering this type of intervention through this digital medium.

### Mindfulness

The mindfulness component of this study will be based on the mindfulness-based stress reduction developed by Jon Kabat-Zinn [69]. This will involve 8 weeks of homebased formal and informal mindfulness practice which will include body scan, sitting, walking, mountain and loving-kindness practice [27,28,70]. Similar to the exercise interventions all mindfulness practices and materials are within the app allowing participants to access these when it suits them. Each week new materials and mindfulness practices will be added to the app allowing participants to build the practice as the weeks progress. The estimated time to complete each weekly online session will be between 1-2 hours with daily practices varying between 10 minutes in the initial weeks to up to 25 mins for 5-6 days per week by week 8, this can be adapted to suit the individual schedule and goals which will be discussed with participants during the weekly contact call. Logging of mindfulness practice will be encouraged through the daily app reminder. All participants will be screened to ensure there are no contraindications to undertaking mindfulness. Even though it is generally well tolerated, there is a possibility of side effects such as increased anxiety, participants will be made aware of this and encouraged to report any issues to the research team [71].

### Digital procedure

Participants will be sent written detailed information on how to download the mobile app. The first step involves registration via a portal which allows the secure collecting of app data. This will generate an e-mail which gives a link to the downloading of the app dependent on the device the participant uses [72]. Once app is downloaded, logged in and opened participants will view a home page which will give a running total of activity/mindfulness material accessed and walking/exercise goals achieved. The home tab also has a send report feature where participants can upload reports of the activity for the day. The other tabs at the top include activity and mindfulness which once clicked on will display the list of material for these weeks and a visual graph which enables self-monitoring. Within the activity tab participants can set their goals and also send reports via this area. The app is designed to take into account randomisation so that participants will only be able to view the content relevant to the group they drew during randomisation. The data entered by the participants will be able to be viewed by the study team via an administration portal allowing for monitoring and feedback to participants [72]. (Fig 3. screen shots of app).

**Fig 3.**
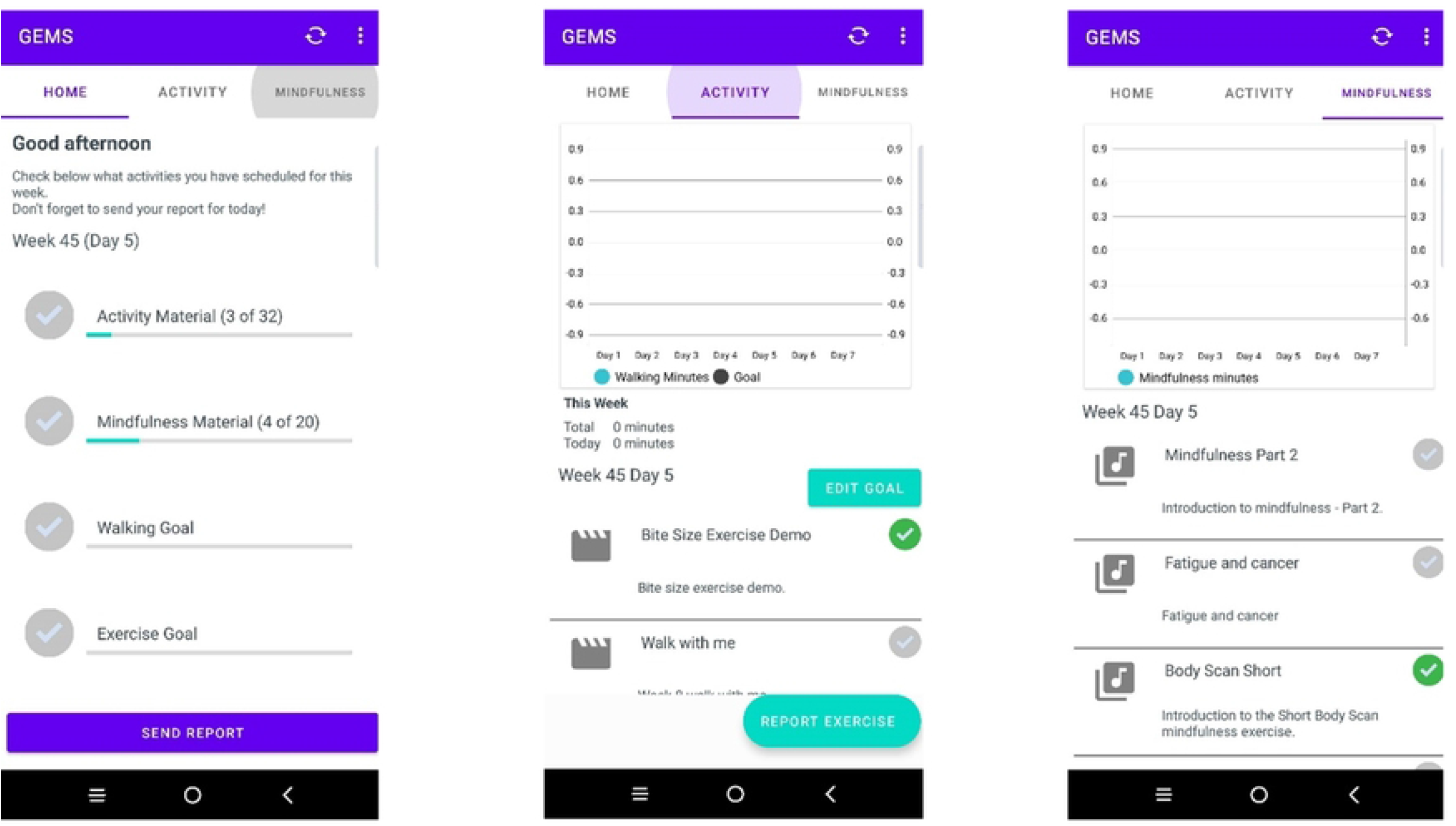
Mobile app screen shots.

### Process evaluation

Following completion of the 8-week intervention, participants will be invited to take part in focus groups to allow evaluation of acceptability and satisfaction of taking part in the study from the participants perspective. As the whole intervention will be conducted remotely, focus groups will be facilitated online via Microsoft Teams. This format for focus group has certain reported advantages, such as reduced dropouts, more relaxing for participants resulting in better communication, cost effectiveness, time saving in travel and the ability to include participants who live in more remote areas from the study centre [73]. The number and size of groups will be dependent on trial recruitment numbers and continue until data saturation, which will be reached when no new information is generated [74]. Separate groups will take place for each intervention group to ensure that each group can share and reflect on their experience of the intervention they took part in. Probe open-ended questions shaped by relevant literature and study objectives will be used to keep the discussion flowing. All focus group discussion will be audio and video recorded, then transcribed verbatim for analysis [75] Focus groups data will be subjected to Thematic Analysis. This type of analysis is flexible in that it allows for the in-depth identification and interpretation of themes [76,77]. The phases of the analysis will include: familiarising with data, generating initial codes, searching for themes, reviewing themes, defining and naming themes and integrating these themes into a final report for publication [78].

### Sample size

The sample size for this study is based upon feasibility and review of similar studies. The literature indicates that feasibility studies of similar interventions and methodologies within the cancer population recruit on average 40 participants in total [57,79,80]. Therefore, it is anticipated that this study will recruit a total of 40 participants with 20 in each arm of the study.

### Statistical analysis

Descriptive statistics will be performed to produce mean values and standard deviations. Groups will be compared at baseline using independent t-test (two independent samples) for continuous data, and the chi-square test for independence for categorical data.

Primary analyses will be performed using the intention to treat on all data and using imputation method where outcomes are missing. Alpha will be set a priori at a level of p= 0.05. Repeated-measures ANOVAs will then be used to determine the effects of exercise and mindfulness and mindfulness intervention on changes in all outcomes of interest.

As this is a feasibility study and preliminary efficacy study, effect sizes (Cohen’s d) will be calculated by taking the mean difference and dividing by the pooled standard deviation to better estimate the meaningfulness of change for each observed outcome following the intervention. The values and meanings for effect size estimates are a small effect size (d=0.2), a moderate effect size (d=0.5), and a large effect size (d=0.8).

Corelation analysis will be performed to determine whether changes in mindfulness/exercise are associated with changes in fatigue, anxiety, depression or sleep.

Adherence will also be assessed via the app through the self-reports of activity that participants complete and usage of the materials. Studies have suggested that dose may have an impact on outcomes with greater adherence showing more favourable outcomes [81]. The relationship between app adherence, dose and outcomes will be explored in this study.

### Data management and protection

Participants will be allocated a unique code so that anonymity is protected. All data will be stored on a secure server at Ulster University. Databases will be encrypted, as will any transferred data and held securely on an repository. The key to the participant IDs will be held by the Chief Investigator. Data will be kept on password-protected computers that can only be accessed by the research team. Access to the anonymised data will use authentication and be traceable by login details. Participants will be made aware that confidentiality will be maintained but the research team will be unable to prevent other members that participate in online focus groups to adhere to this confidentiality agreement. Participants will also be informed that if they withdraw from this part of the study that once data has been anonymised it may be difficult to remove it.

### Ethics

Ethical approval has been granted from Ulster University Research Ethics Committee (REC.21.0076).

### Dissemination Policy

The findings of the research will be communicated with those who participated in the study either via email or a local event. The researcher will disseminate the projects findings and outputs via oral and poster presentations at local, national and international conferences and publications in peer reviewed journal papers.

## Discussion

Fatigue continues to be a distressing symptom for many cancer patients either pre diagnosis, diagnosis, during and post treatment. For some it continues to be a long term symptom that effects all aspects of life including social, biological, financial and emotionally leading to reduced quality of life. The current evidenced based advice and interventions offered for managing this symptom include exercise, nutrition advice, sleep hygiene, or prioritising, pacing and planning activities [82]. In recent years other interventions such as mindfulness, have been shown to have a favourable responses for CRF, which in turn also demonstrated a positive effect on stress and sleep all of which are identified as part of the cluster symptom for fatigue [83]. However, there appears to be little research that examine the use of more than one intervention simultaneously. This study will not only explore whether this is feasible but also if the simultaneous delivery of interventions results in any additionally effect. Furthermore, the use of an active comparator will be of value, as it has been suggested previously that a usual care or wait list control can result in a greater result in the experimental arm [35]. Additionally, it has been indicated that assessing the levels of fatigue prior to entering a study can have an impact on the effectiveness of the intervention. This was demonstrated in a meta-analysis that examined the effectiveness of physical activity on fatigue for colorectal cancer patients. It found that levels of fatigue in some of the included studies within this review were equivalent to the normal population which resulted in participant outcome measures showing little improvement therefore described as a ceiling effect for the interventions [42]. Therefore, this study has been designed around these concepts and will be both assessing levels of fatigue as part of the screening process and incorporating mindfulness as the active comparator arm.

This study will also use a digital platform for delivering this intervention. By exploring the feasibility of delivering the intervention through this platform it may also make it feasible in the future to diversify this app and its delivery for other types of cancer populations and possibly management of other symptoms.

### Trial registration data

Primary registry and trial identifying number:NCT05561413

Date of registration in primary registry: September 20 2022

Secondary identifying numbers: CN-02467186

Source of monetary or material support: Department for the Economy Northern Ireland

Primary sponsor: Ulster University

Contact for public enquires: Dr Jackie Gracey

Contact for scientific enquires: Professor Ciara Hughes

Public Title: Gynaecology Exercise and Mindfulness Study (GEMS)

Scientific title: Randomised Controlled Feasibility Trial Evaluating the Effectiveness of Mindfulness Compared to Exercise and Mindfulness on Fatigue in Women with Gynaecology Cancer

Countries of recruitment: UK

Health condition or problems: Gynaecology cancer, cancer related fatigue

Intervention: Intervention: Mindfulness and exercise Active comparator: Mindfulness

Key inclusion and exclusion criteria: Ages eligible for study: > 18 years Inclusion: Women diagnosed with gynaecology cancer, still experiencing fatigue > 4 on NRS scale Exclusion: Currently practicing mindfulness, diagnosis of schizophrenia, bipolar, psychosis risk, existing medical condition inhibits safe participation.

Study type: Interventional. Allocation: Randomised parallel 1:1 Primary purpose: feasibility Date of first enrolment: June 2022

Target sample size: 40

Recruitment status: Recruiting

Primary outcomes: Feasibility: eligibility, retention, attrition and adherence

Key Secondary outcomes: Fatigue, quality of life, sleep, psychological outcome: anxiety, depression, mindfulness and physical activity

## Data Availability

No datasets were generated or analyses during the current study. All relevant data from this study will be made available upon study completion.

## Author Contributions

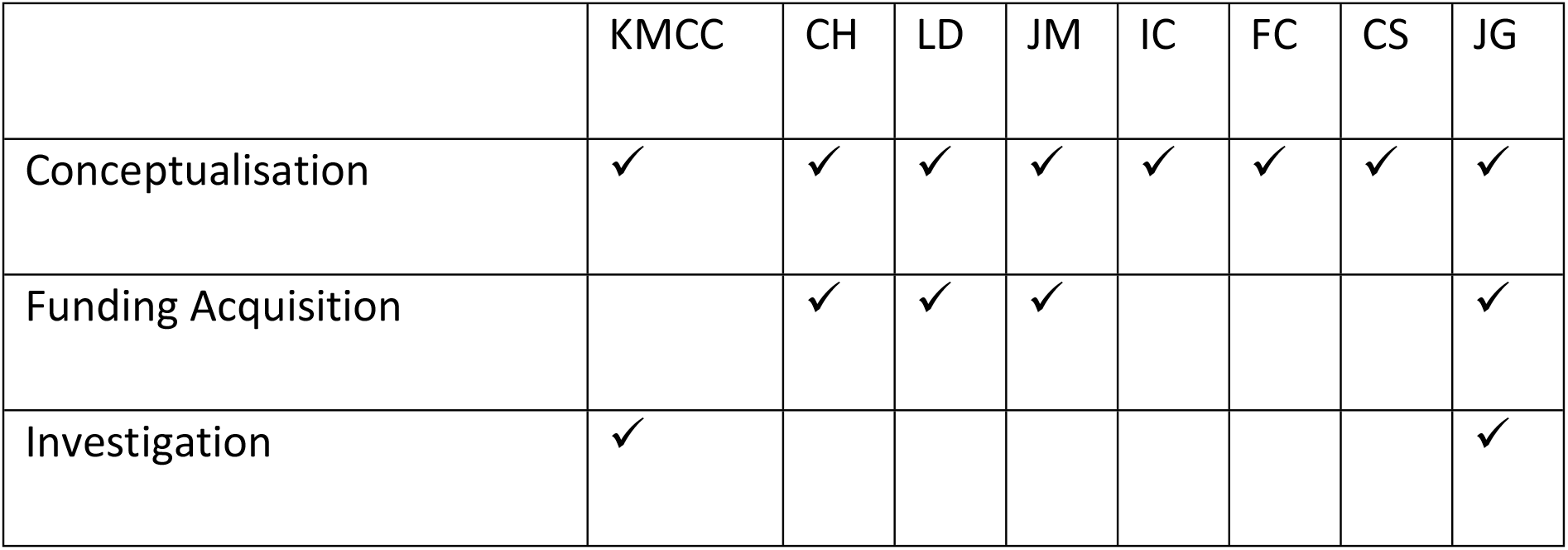
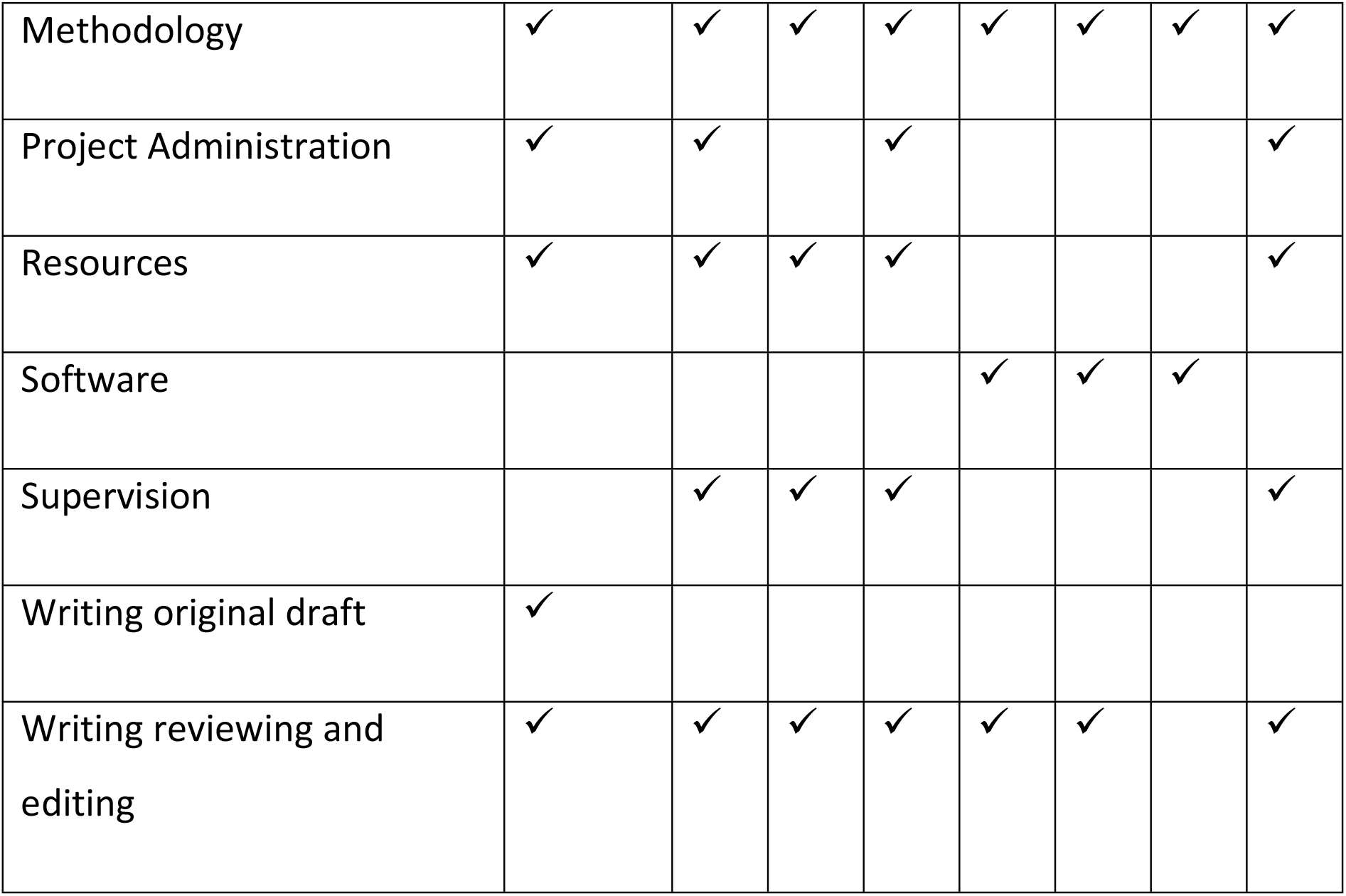

